# Pre-infection 25-hydroxyvitamin D3 levels and association with severity of COVID-19 illness

**DOI:** 10.1101/2021.06.04.21258358

**Authors:** Amiel A. Dror, Nicole G. Morozov, Amani Daoud, Yoav Namir, Yakir Orly, Yair Shachar, Mark Lifshitz, Ella Segal, Lior Fischer, Matti Mizrachi, Netanel Eisenbach, Doaa Rayan, Maayan Gruber, Amir Bashkin, Edward Kaykov, Masad Barhoum, Michael Edelstein, Eyal Sela

## Abstract

**Objective:** Studies have demonstrated a potential link between low vitamin D levels and both an increased risk of infection with SARS-CoV-2 and poorer clinical outcomes but have not established temporality. This retrospective study examined if, and to what degree, a relationship exists between pre-infection serum vitamin D levels and disease severity and mortality of SARS-CoV-19.

**Design and patients:** The records of individuals admitted between April 7^th^, 2020 and February 4^th^, 2021 to the Galilee Medical Center (GMC) in Nahariya, Israel with positive polymerase chain reaction (PCR) tests for SARS-CoV-2 were searched for vitamin D (VitD) levels measured 14 to 730 days prior to the positive PCR test.

**Measurements:** Patients admitted to GMC with COVID-19 were categorized according to disease severity and VitD level. Association between pre-infection VitD levels and COVID-19 severity was ascertained utilizing a multivariate regression analysis.

**Results:** Of 1176 patients admitted, 253 had VitD levels prior to COVID-19 infection. Compared with mildly or moderately diseased patients, those with severe or critical COVID-19 disease were more likely to have pre-infection vitamin D deficiency of less than 20 ng/mL (OR=14.30, 95%, 4.01-50.9; p < .001); be older (OR=1.039 for each year, 95% CI for OR, 1.017-1.061; p< .01), and have diabetes (OR=2.031, 95% CI for OR, 1.04-3.36; p= 0.038). Vitamin D deficiency was associated with higher rates of mortality (p<0.001) and comorbidities including COPD (p=0.006), diabetes (p=0.026), and hypertension (p=0.016).

**Conclusions:** Among hospitalized COVID-19 patients, pre-infection deficiency of vitamin D was associated with increased disease severity and mortality.

## Introduction

Vitamin D (VitD) is most often recognized for its role in bone health, but low VitD levels have been associated with a range of autoimmune, cardiovascular, and infectious diseases due to its role as an essential immunologic mediator ^1^. Experimental laboratory evidence evaluating the impact of VitD on immunological responses has shown inhibitory effects on the production of pro-inflammatory cytokines, including TNF-alpha and IL-6, by various mechanisms, including down-regulating viral-induced NFkB activation ^2^.

Vitamin D deficiency (VDD) is a global health problem, and its high prevalence in the Middle East is well established ^3^. Epidemiological risk factors for VDD include greater skin pigmentation, low sun exposure, use of skin-covering clothes, and a diet low in fish and dairy products. Studies have previously demonstrated that social habits in specific ethnic groups and a preference to wear long clothing outdoors are independent risk factors for VDD, particularly among women ^4^. Consistent with laboratory data, clinical studies have established an association between low vitamin D levels and an increased risk of acquiring influenza and respiratory viruses ^5^. Meta-analyses of randomized controlled trials conducted between 2007 and 2020 suggest that vitamin D supplementation was safe and reduced the risk of acute respiratory infection compared with placebo ^6^.

COVID-19 is an acute respiratory infection caused by the SARS-CoV-2 virus, which emerged in China in December 2019 and rapidly developed into a global pandemic. Factors associated with poorer COVID-19 prognoses include geographic location in northern countries, older age, darker skin pigmentation, BAME ethnicity (Black and minority ethnic groups), male sex, obesity, and preexisting conditions such as diabetes and hypertension; these risk factors are also independently associated with VDD ^7^.

As with other respiratory infections, a link between VDD and COVID-19 infection is emerging ^8,9^. Low serum 25-hydroxyvitamin levels among hospitalized COVID-19 patients have also been linked with increased disease severity and poorer clinical outcomes ^10^. In addition, hospitalized COVID-19 patients have been shown to present lower mean and median levels of VitD than the general population and COVID-19 outpatients ^11^. However, serum VitD levels are often measured during hospitalization for COVID-19. When this is the case, determining the direction and temporality of the association between acute COVID-19 disease and low VitD serum levels is a challenge. In other words, it is difficult to ascertain a definitive causative effect of baseline VitD level on a clinical presentation during active COVID-19 infection.

To better understand the temporal sequence between low VitD levels and association with severity of acute COVID-19 disease course, we determined whether the severity of disease among patients admitted with acute COVID-19 correlated with their most recent pre-infection VitD serum levels.

## Methods

### 1. Participants

This was a retrospective study. Prior to study initiation, ethical approval was granted by the Research Ethics Committee of the Galilee Medical Center 0204-20-NHR. Inclusion criteria were adult patients with PCR-confirmed COVID-19 infection admitted to Galilee Medical Center (GMC) between April 7^th^, 2020, and February 4^th^, 2021. Patients under the age of 18 years old were excluded from this study.

### 2. Measurement

COVID-19 infection was confirmed by two independent positive polymerase chain reaction (PCR) tests. Clinical data collected during the inpatient hospital stay at Galilee Medical Center included length of hospitalization in days, the severity of illness (critical, severe, moderate, and mild, as determined by the WHO definition of COVID-19 disease severity (WHO/2019-nCoV/clinical/2020.5)), mortality during hospitalization, and comorbidities (COPD, chronic heart failure, chronic ischemic heart disease, chronic renal disease, diabetes, and hypertension). Additional demographic characteristics included age, ethnicity, and BMI (Table 1).

**Table 1.** Characteristics of Covid-19 hospitalized patients stratified by most recent <u>vitamin D</u> level from 14 to 730 days before COVID-19 test

| Characteristics | No. (%) |  |  |  | *P*-value*<br>2-sided<br>(between<br>groups) | *P*-value*<br>2-sided<br>(<20<br>compared<br>with ≥20) |
| --- | --- | --- | --- | --- | --- | --- |
|  | Most recent vitamin D level (ng/mL) before COVID-19 test<br>(N=253) |  |  |  |  |  |
|  | <20 | 20-29.9 | 30-39.9 | ≥40 |  |  |
| No. of patients | 133 | 36 | 44 | 40 |  |  |
| Age, y |  |  |  |  |  |  |
| mean (std) | 68.4 (15.9) | 62.2 (17.9) | 55.7 (20.7) | 57.7 (21.0) | <0.001§ |  |
| Range |  |  |  |  |  |  |
| <50 | 18 (13.5) | 8 (22.2) | 18 (40.9) | 17 (42.5) | <0.001§ | <0.001 |
| 50-65 | 36 (27.1) | 14 (38.9) | 10 (22.7) | 6 (15.0) |  |  |
| ≥65 | 79 (59.4) | 14 (38.9) | 16 (36.4) | 17 (42.5) |  |  |
| Sex |  |  |  |  |  |  |
| Female | 69 (51.9) | 18 (50.0) | 30 (68.2) | 27 (67.5) | 0.102 | 0.099 |
| Male | 64 (48.1) | 18 (50.0) | 14 (31.8) | 13 (32.5) |  |  |
| Ethnicity |  |  |  |  |  |  |
| Arabs | 83 (64.3) | 16 (44.4) | 33 (76.7) | 19 (47.5) | 0.006 | .298 |
| Non-Arabs | 46 (35.7) | 20 (55.6) | 10 (23.3) | 21 (52.5) |  |  |
| COVID-19 disease severity category (WHO) |  |  |  |  |  |  |
| Mild | 7 (5.3) | 8 (22.2) | 37 (84.1) | 28 (70.0) | <0.001 § | <0.001 § |
| Moderate | 50 (37.6) | 24 (66.7) | 3 (6.8) | 9 (22.5) |  |  |
| Severe | 64 (48.1) | 3 (8.3) | 4 (9.1) | 3 (7.5) |  |  |
| Critical | 12 (9.0) | 1 (2.8) | 0 (0.0) | 0 (0.0) |  |  |
| BMI |  |  |  |  |  |  |
| mean (std) |  |  |  |  |  |  |
| <30 | 98 (73.7) | 32 (88.9) | 34 (77.3) | 34 (85.0) | 0.163 | 0.069 |
| >30 | 35 (26.3) | 4 (11.1) | 10 (22.7) | 6 (15.0) |  |  |
| Mortality during hospitalization |  |  |  |  |  |  |
| Yes | 34 (25.6) | 1 (2.8) | 1 (2.3) | 2 (5.0) | <0.001 | <0.001 |
| No | 99 (74.4) | 35 (97.2) | 43 (97.7) | 38 (95.0) |  |  |

| Comorbidities |  |  |  |  |  |  |
| --- | --- | --- | --- | --- | --- | --- |
| COPD | 82 (61.7) | 15 (41.7) | 20 (45.5) | 18 (45.0) | .048 | 0.006 |
| Chronic Heart failure | 16 (12.0) | 3 (8.3) | 1 (2.3) | 2 (5.0) | 0.211‡ | .072 |
| Chronic Ischemic Heart Disease | 16 (12.0) | 3 (8.3) | 3 (6.8) | 1 (2.5) | 0.316‡ | .124 |
| Chronic renal disease | 17 (12.8) | 5 (13.9) | 3 (6.8) | 3 (7.5) | .604‡ | .425 |
| Diabetes | 56 (42.1) | 9 (25.0) | 12 (27.3) | 13 (32.5) | .125 | .026 |
| Hypertension | 82 (61.7) | 16 (44.4) | 21 (47.7) | 18 (45.0) | .092 | .016 |

Abbreviations: BMI, body mass index (calculated as weight in kilograms divided by height in meters squared); COVID-19, coronavirus disease 2019; Patient data includes COVID-19 admission between Apr 7<sup>th</sup>, 2020 to Feb 4<sup>th</sup>, 2021 who had serum vitamin D level measured between two years to one month before admission. Most recent Vitamin D level was depicted for the purpose of the study.
\* P values were determined using the Pearson Chi-Square test unless mentioned otherwise.
‡ P values were determined using the Fisher-Freeman-Halton Exact Test
§ P values were determined using Kruskal Wallis Test
† P values were determined using Mann whitney Test

Disease severity was calculated at the point of highest severity during the patient’s inpatient stay; as an example, a patient who arrived at the hospital in mild condition but decompensated to a critical condition during admission would be classified as having had a critical illness. The COVID-19 inpatient group with historical VitD levels admitted to Galilee Medical Center is herein referred to as the “VitD group.” The VitD group’s 25(OH)D levels were obtained from their respective electronic medical records 14 to 730 days prior to COVID-19 diagnosis. The most recent VitD level from prior to infection with COVID-19 was utilized. Patients with previous VitD measurements had likely been tested either as part of a routine blood workup or following a clinical suspicion for VDD. We did not have access to data regarding whether patients were treated for VDD. Patients’ 25(OH)D levels were divided into four universally accepted categories: deficient (below 50 nmol/L or 20 ng/ml), insufficient (50 nmol/L to 75 nmol/L or 20 -29.9 ng/ml), adequate (75-99.75 nmol/L or 30-39.9 ng/ml), and high-normal (above 187.5 nmol/L or 40 ng/mL).

The data utilized for this study is available upon request from the corresponding author.

### 1. Analysis

Continuous variables were presented as the mean ± standard deviation. Comparisons of continuous variables among groups were examined with either the ANOVA, Kruskal Wallis test, independent sample t-test, or Mann-Whitney test. Those tests were chosen according to the number of the compared groups, the sample size of the groups, and the variables’ distribution. Ordinal variables were compared among the groups with the Kruskal Wallis test or the Mann-Whitney test and categorical variables using the Pearson’s chi-square or the Fisher’s exact test (if expectancy<5). With the intention to complete the univariate comparisons of the characteristics between patients, we examined the relationships among several potential predictors of COVID-19 disease severity and the disease severity degree itself. Finally, we examined the correlation between the VitD level and the disease severity degree, adjusted for independent variables which were found to be significant in the univariate analysis or according to theoretical consideration such as age and gender. By calculating the odds ratio (OR) along with 95% confidence intervals (CI), using a multivariate logistic regression model (with the Backward elimination method). We used a statistical significance threshold of P<0.05. The analysis was performed using the IBM SPSS Statistic software, version 27.0.

## Results

Of 1176 individuals admitted for COVID-19 at the Galilee Medical Center between April 7^th^, 2020, and February 4^th^, 2021, historical VitD levels were obtained between 14 and 730 days before the first positive COVID-19 test for 253 (21.5%) individuals (mean age 63.3 [SD=18.6] years; 144 [56.9%] women). Of the 253 individuals with pre-infection VitD levels, 133 (52.5%) had a level less than 20 ng/mL, 36 (14.2%) had 20 to less than 30 ng/mL, 44 (17.3%) had 30 to less than 40 ng/mL, and 40 (15.8%) had 40 ng/mL or greater (Table 1). Mortality among patients with sufficient VitD levels was 2.3%, in contrast to the VitD deficient group’s 25.6% mortality rate (p-value<0.001) (Table 1). Stratified by COVID-19 disease severity, lower vitamin D levels were more common in patients with severe or critical disease (<20 ng/mL: 76 of 87 individuals [87.4%]) than individuals with mild or moderate disease (<20 ng/mL: 57 of 166 individuals [34.3%] p-value<.001) (Table 2).

**Table 2.**
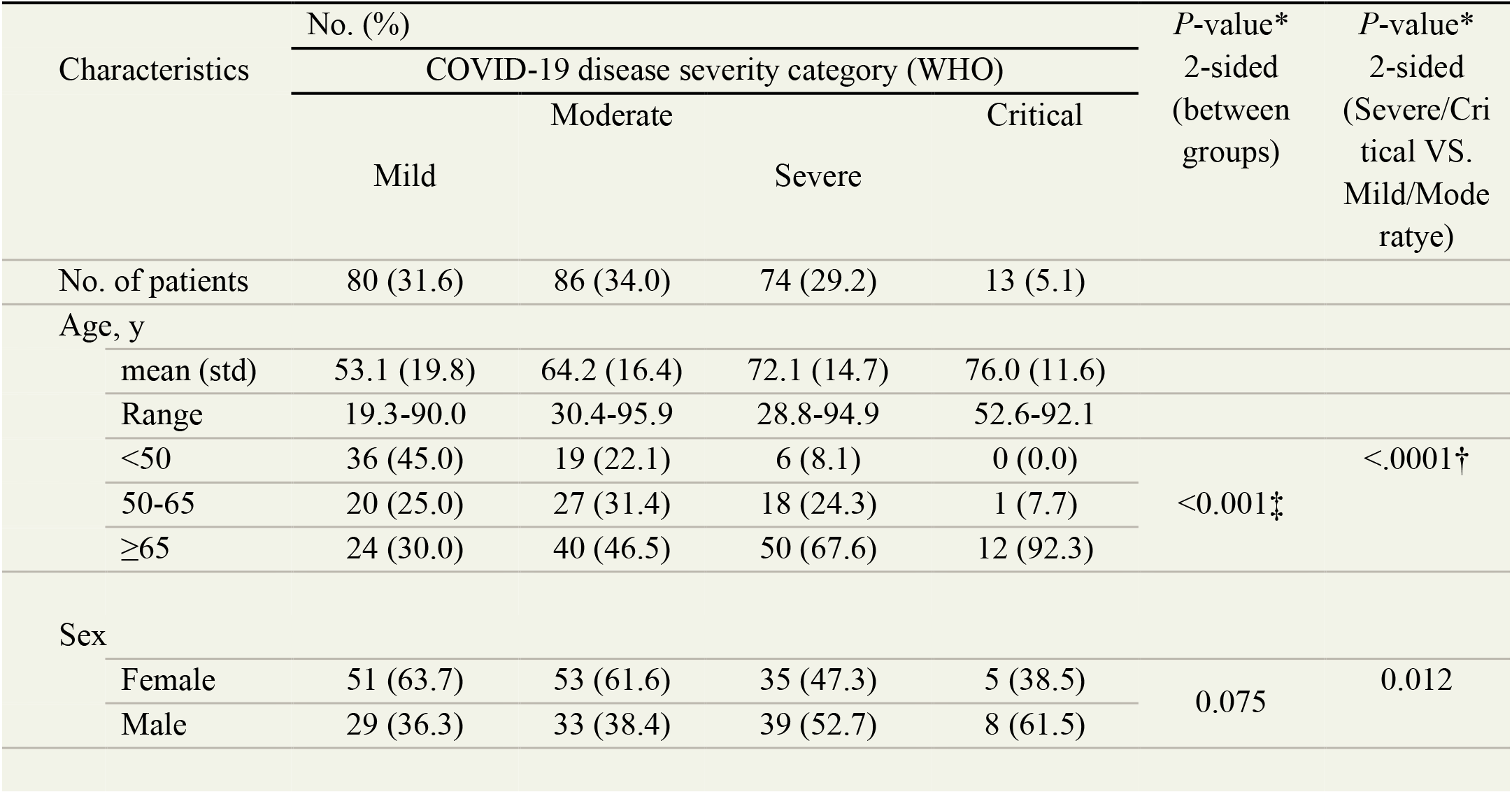
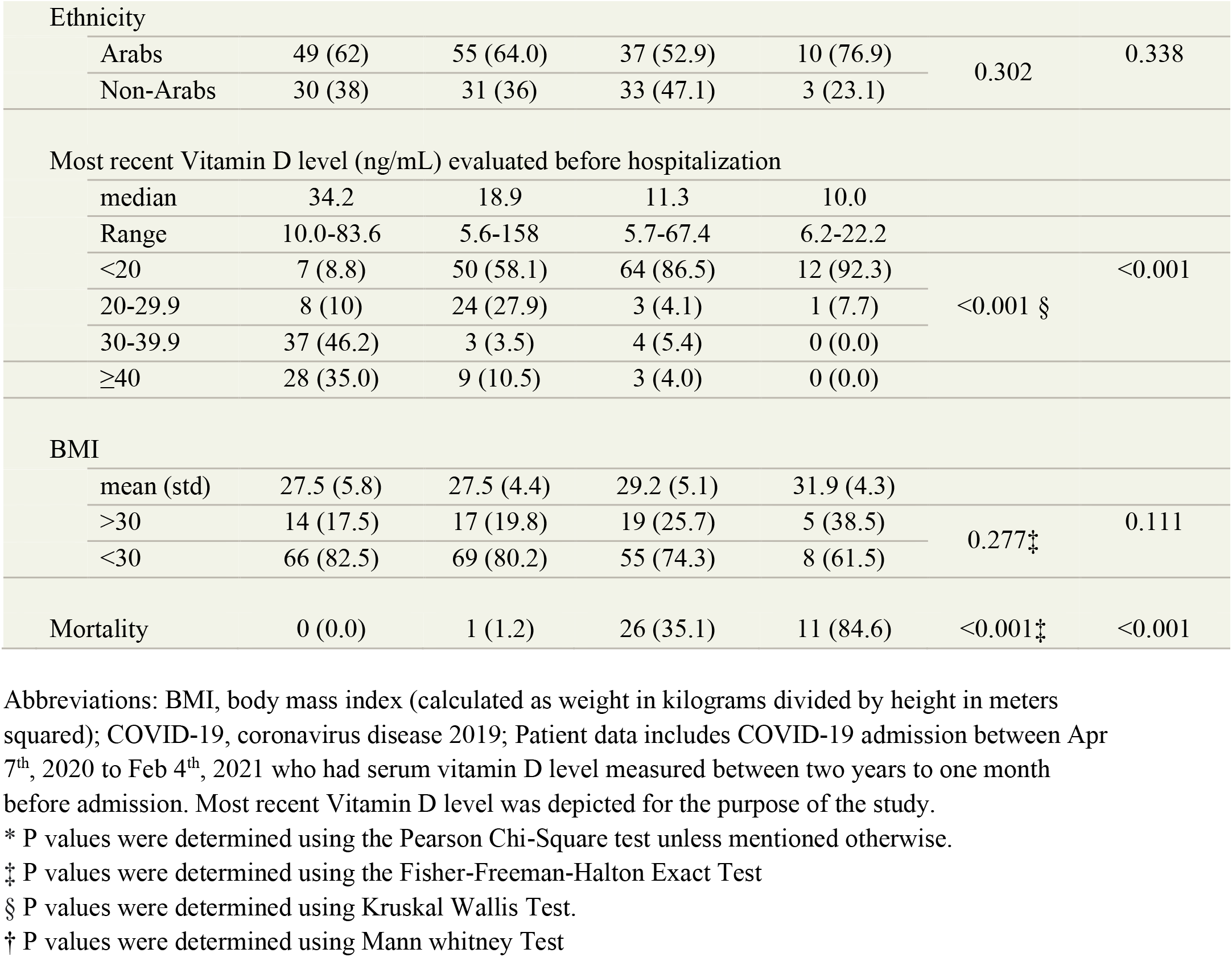
Characteristics of Covid-19 hospitalized population with recent <u>vitamin D</u> level stratified by COVID-19 disease severity category

Multivariate analysis in individuals with prior VitD levels found that compared with those with mild or moderate COVID19 disease, those with severe or critical disease were14 times more likely to have VitD deficiency <20g/nl (OR=14.30, 95% CI, 4.01-50.9; p-value < .001) (Table 3). Multivariate analysis in individuals with prior VitD levels found that compared with those with mild or moderate COVID19 disease, those with severe or critical disease were14times more likely to have VitD deficiency <20g/nl (OR=14.30, 95% CI, 4.01-50.9; p-value < .001) (Table 3). The variables included in step 1 of this model were age, gender (male), BMI, and comorbidities. The final model represents the last step of the backward elimination process following the adjustment of confounding factors (Table 3).

**Table 3.** Multivariate logistic regression analysis of possible predictors of **severe or critical** covid-19 disease among hospitalized patients with recent <u>vitamin D</u> level from 14 to 730 days before COVID-19 test - **Last step in the Logistic regression (backward elimination**)

|  |  |  |  |  |  |  |  |
| --- | --- | --- | --- | --- | --- | --- | --- |
|  | Mild/moderate cases |  | Severe/Critical cases |  | OR | 95%CI | P value |
| variable | n | % | N | % |  |  |  |
| Vit D levels |  |  |  |  |  |  |  |
| ≥40 | 37 | 92.5% | 3 | 7.5% | baseline |  |  |
| 30-39.9 | 40 | 90.9% | 4 | 11.1% | 1.450 | 0.290- 7.249 | 0.651 |
| 20-29.9 | 32 | 88.9% | 4 | 11.1% | 1.315 | 0.255- 6.780 | 0.743 |
| <20 | 57 | 42.9% | 76 | 57.1% | 14.308 | 4.016- 50.974 | 0.000 |
| Mean Age, y (std) | 58.9 (18.9) |  | 72.7 (14.3) |  | 1.039 | 1.017- 1.061 | 0.001 |
| Diabetic |  |  |  |  |  |  |  |
| yes | 45 | 48.3% | 42 | 51.7% | 2.031 | 1.041- 3.965 | .038 |
| no | 121 | 72.9% | 45 | 27.1% | baseline |  |  |
| Constant |  |  |  |  | .004 |  | .000 |

Multivariate Logistic regression analysis, Backward elimination method. Dependent variable: Covid-19 disease severity (severe or critical vs. mild or moderate). The variable included in the step 1 of this model: Age, Gender (male), BMI, Chronic obstructive pulmonary disease (COPD), Chronic Heart failure (CHF), Chronic Ischemic Heart Disease (CIHD), Chronic renal failure (CRF), Diabetes and VitD level. OR, Odds ratio; CI, Confidence interval.

A comparison of VitD levels among COVID-19 disease severity categories demonstrated a progressive decrease in levels of VitD as the disease severity increased (Figure 1). A significant difference in VitD level was found between mild compared with moderate disease categories (P =0 .001) and moderate compared with severe (P =0.002) (in the mild group, 8.8% had a level less than 20 ng/mL, as well as 58.1% in the moderate group, and 86.5% in the severe patients) (Figure 1 and Table 2). No difference between severe and critical individuals with regards to VitD (P =0.405) was observed (Figure 1).

**Figure 1.**
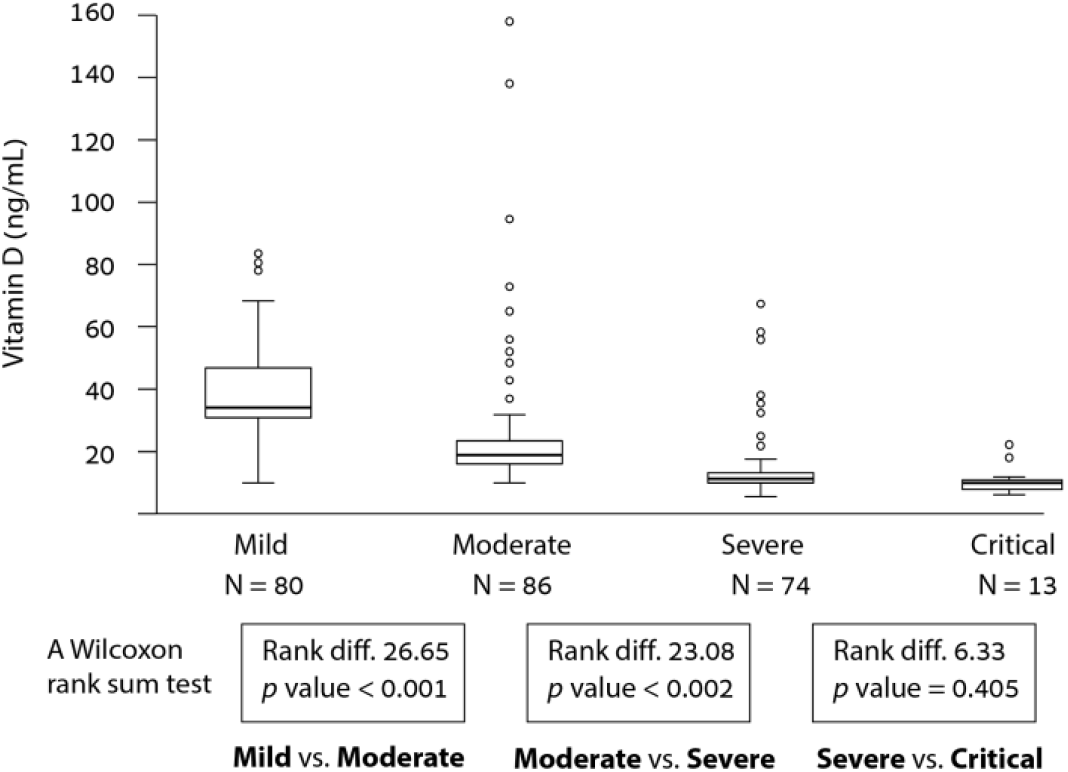
Box-and-whisker plot of most recent serum 25-hydroxyvitamin D levels before hospitalization were collected as a baseline (N=253). The mean vitamin level compared between the four categories of COVID-19 disease severity as determined by the WHO definition (WHO/2019-nCoV/clinical/2020.5). A Kruskal-Wallis-test for multiple categories comparison show significant difference between groups *p*-value <0.001. A Wilcoxon rank sum test compared vitamin D mean rank of two neighboring category sequentially; Mild compared with Moderate (mean difference, 12.96 ng/mL; [Rank difference 26.65] P < .001); Moderate compared with Severe (mean difference, 10.72 ng/mL [Rank difference 23.08]; P < .002); Severe compared with Critical (mean difference, 3.96 ng/mL [Rank difference 6.33]; P =0.405). The boxes represent the range of vitamin D values within the interquartile range (50% of the cases). The whiskers outside the box mark the most upper and lower values within 1.5 times the interquartile range. Outliers’ values in each group represented with empty circles.

While both age and VitD baseline are independent predictors of increased disease severity, further analysis found a significant correlation between VDD when stratifying patients into three age groups (<50, 50-65, ≥65) (Figure 2). The strongest correlation between lower VitD level and COVID-19 disease severity was seen in ≥65 years old patients (r =-0.717; p-value < 0.001). In patients under 50 years of age, COVID-19 severity was still correlated with VitD deficiency but to a lesser extent (r =-0.566; p-value < 0.001).

**Figure 2.**
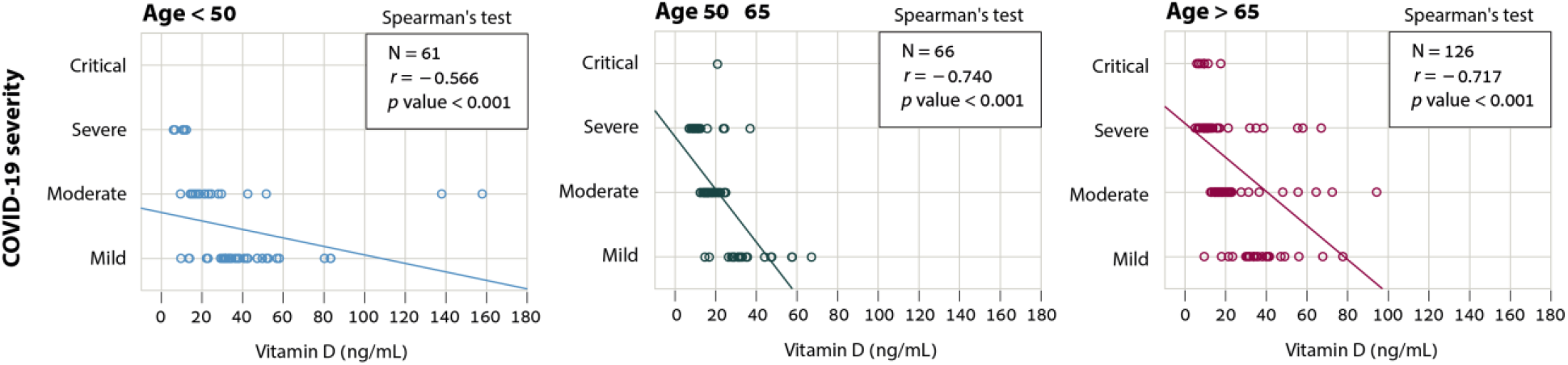
Correlation between VDD and COVID-19 disease severity stratified by three different group of age (<50, 50-65, ≥65). The severity of illness (critical, severe, moderate, and mild, as determined by the WHO definition of COVID-19 disease severity (WHO/2019-nCoV/clinical/2020.5))

## Discussion

In this retrospective single-institution study, we demonstrate a correlation between low vitamin D levels prior to COVID-19 infection and increased COVID-19 disease severity and mortality during hospitalization.

The vital role of VitD during the COVID-19 pandemic has been highlighted beyond its well-known benefit for bone and muscle health as a regulatory component of the innate and adaptive immune response during viral infections. Other hallmarks of COVID-19 disease have been linked with VitD status, including the cytokine storm that portends acute respiratory distress syndrome. Several studies demonstrated the association of VDD in a patient’s history with increased risk for positive COVID-19 test results ^12,13^. Existing research has suggested a correlation between low 25-hydroxyvitamin D levels upon admission as a predictor of poorer COVID-19 disease outcomes ^14,15^. However, a low level of serum VitD measured during the acute COVID-19 infection may reflect a consequence of chronic inflammation rather than an underlying cause. By using the most recent historical VitD levels, the obtained vitamin D value in our study assuredly preceded COVID-19 infection.

We found that lower VitD levels were associated with greater disease severity and mortality. While 48.1% of VitD deficient <20 ng/mL patients had severe disease courses, less than 10% of patients with VitD levels ≥20 ng/mL had severe courses. While the mortality of patients who had VitD levels ≥20 ng/mL was at 5% or lower, the mortality of VitD deficient patients <20 ng/mL was much higher, at 25.6%. Older age is associated with both VitD deficiency and poorer COVID-19 outcomes. To adjust for this variable, we performed a multivariate analysis which adjusted for age as a confounder, demonstrating that pre-infection VDD increased the risk of severe COVID-19 disease, independent of age. Reduced VitD levels in younger COVID-19 inpatients may suggest an increased hospitalization risk due to COVID-19 illness in this group with severely low VitD while older patients remain at increased risk of hospitalization for COVID-19, even at higher levels of the VDD range.

The higher ratio of female-to-male patients in the VitD group (56.9% females) compared to the non-VitD group (50.4% females) may be explained by the higher risk of osteoporosis, and subsequently vitamin D testing, in women. Status as a postmenopausal woman is a known risk factor for the development of VDD, thus increasing the likelihood of a woman having had VitD testing ^16^. Because VitD and calcium supplementation have been shown to reduce the risk of fractures in the elderly, there is the possibility that patients with previously low VitD results have taken supplementation, which significantly increases serum VitD levels. Furthermore, previous studies have established that Israel, a sunny country, has high levels of VitD insufficiency or deficiency; this may predispose individuals or their providers to consider supplementation, particularly for women ^17^.

Despite the significant propensity for VDD among Arab ethnicities, our data showed no effect of ethnicity on disease severity and mortality (Tables 1 and 2). This evidence implies that baseline serum VitD level is more significant to COVID-19 disease course than ethnicity, at least in our cohort. While the high percentage of VDD in Arabs (64.3% of patients with VitD <20 ng/mL) compared with others was consistent with previous research demonstrating high rates of VDD in Arabs, including in Arab-American and Arab-European populations, this may be the result of a higher frequency of VitD testing in this population ^18,19^. A proposed explanation for this finding is a higher level of skin pigmentation that decreases dermal vitamin D synthesis ^20^. It is not unlikely that the skin’s increased pigmentation, combined with a conservative dress, are partly responsible for these findings. Other mechanisms for VitD imbalances among different ethnic groups are subject to investigation and span across inherent medical factors and social and economic considerations of health ^21^. Preexisting medical conditions, healthcare access, and other factors may contribute to the disproportionate impact of COVID-19 on minority populations ^22^. Interestingly, while high VitD levels (40 ng/mL) among Black individuals decrease the risk for COVID-19, high VitD levels in ethnic minority groups in Israel showed no benefit of lower COVID-19 disease severity compared with a sufficient VitD level ^13^.

There are several important limitations of the study. First, vitamin D deficiency can be a part of a wide range of chronic health conditions or behavioral factors that simultaneously increase COVID-19 disease severity and mortality risks. As an example, COPD, while associated with VDD in our study, is a known risk factor for poorer COVID-19 outcomes. It should be clearly stated that a patient’s historically low VitD levels do not cause COVID-19 but rather may increase the likelihood of SARS-COV-2 infection and morbidity and mortality under various clinical settings ^23–25^. Moreover, VitD correction in the community setting preceding the infection will not necessarily improve the future COVID-19 disease course of an affected individual. Patients’ supplementation history was not obtained or analyzed as part of this research. The use of historical results from community health providers may be influenced by prior VDD correction therapy given due to low serum levels, the effect of which is difficult to fully deduce.

However, our cohort’s strong correlation between prior VDD and COVID-19 disease outcome implies that most patients remain with low VitD levels when contracting COVID-19 infection. Third, despite our findings, it must be acknowledged recent study in India found no association between VDD and increased mortality and morbidity in hospitalized patients ^26^. In this prospective study in which patients’ VitD levels were measured during hospitalization, the patients had a median age of 54, lower than our cohort’s median age of 63.6; as mentioned, older age is associated with increased mortality due to COVID-19.

Additional confounders that possibly contribute to COVID-19 outcome differences between published cohorts stem from the genetic heterogenicity of different populations ^21^. A panoply of studies highlights the role of host genetics in susceptibility for COVID-19 disease via polymorphism of different key alleles such as ACE, TMPRSS2, Vitamin D binding protein, and other protein-coding genes which participating in the human innate immune system ^27–29^.

Our results add to the existing literature suggesting a potential protective effect of adequate VitD levels in patients with active COVID-19 infection. While our findings have identified an association between pre-infection VDD and COVID-19 severity, these results do not necessarily imply that VitD treatment will impact COVID-19 outcomes, and we should remain cautious about overestimating the potential benefit of VitD supplementation in improving outcomes of SARS-CoV-2 infection. Evidence from a randomized clinical trial involving 240 hospitalized patients with moderate to severe COVID-19 disease showed no benefit of a single high dose of VitD treatment in reducing hospital length of stay ^30^. The relatively long time from symptom onset to randomization (mean of 10.3 days) and VitD treatment in this study limits the extrapolation of the effect of VitD supplementation during early COVID-19 infection. A recently published meta-analysis of 43 randomized controlled trials (RCTs) analyzing data obtained from 48,488 patients showed that vitamin D supplementation was safe and overall reduced the risk of acute respiratory infections compared with a placebo, although the relevance of these findings to COVID-19 is not yet established ^31^. In light of accumulating research on VitD and COVID-19, the US Preventive Services Task Force (USPSTF) released an evidence report concluding that the overall evidence on the benefits of adult screening for vitamin D deficiency is lacking ^32^. Nevertheless, an App-based community survey among 372,720 UK participants showed that dietary supplements, including VitD, lowered the risk of testing positive for SARS-CoV-2 in women but not in men ^33^. Though there is limited data regarding the preventive potential of normal VitD levels, the National Institute for Clinical Excellence (NICE) specifically stresses the importance of regular VitD supplementation during the COVID-19 pandemic, as on ordinary days ^34^.

It should be emphasized that VitD represents only one piece of the complex puzzle that is COVID-19 in addition to underlying comorbidities, genetic predispositions, dietary habits, and geographic factors. Despite the numerous studies related to VitD and COVID-19, to date, there is no evidence supporting the administration of supplementary vitamin D for the treatment of active COVID-19 infection.

## Conclusion

From the early stages of the COVID-19 pandemic, establishing vitamin D deficiency as a risk factor was the aim of many investigators. It was subject to much debate in the general public and multiple medical journals ^35^. Our study contributes to a continually evolving body of evidence that suggests a patient’s history of VDD is a predictive risk factor associated with poorer COVID-19 clinical disease course and mortality. The use of historical results obtained before the COVID-19 pandemic as part of a public health survey enabled us to suggest VDD contributes to the causal pathway of COVID-19 mortality risk and disease severity. Our study warrants further studies investigating if and when VitD supplementation among VitD deficient individuals in the community impacts the outcome of an eventual COVID-19 episode.

## Data Availability

Data is available upon request from the corresponding author.

